# Optimal mobility restriction minimizing COVID-19 and excess suicide deaths in Japan

**DOI:** 10.1101/2021.02.28.21252644

**Authors:** Junko Kurita, Tamie Sugawara, Yoshiyuki Sugishita, Yasushi Ohkusa

## Abstract

**Background:** Strict countermeasures for COVID-19 outbreak such as lockdowns and voluntary restrictions against going out might have reduced mortality because of COVID-19 directly, but might have raised suicide rates.

**Object:** We examined best policies for minimizing overall mortality attributable to COVID-19 directly, and excess mortality by suicide because of COVID-19.

**Method:** We regressed the estimated excess mortality attributable to suicide deaths against mobility-restrictive measures. Mortality attributable to COVID-19 directly was estimated through association between the effective reproduction number and mobility. We sought the best mobility restriction for minimizing overall deaths.

**Results:** Significant association was found between mobility and suicide, but the data were very few. Results showed the best mobility level as 65.5, which represents a 34.5% reduction in mobility from the normal level.

**Discussion and Conclusion:** An overly restrictive policy inducing lower than optimal mobility led to higher total mortality.

## 1. Introduction

Since the emergence of COVID-19, morbidity and mortality have been very low in Japan. As of February 11, 2021, the incidence rate was 3.416 per thousand population. The mortality rate was 0.06 per thousand population [1]. In fact, excess mortality from all causes has remained low in Japan [2]. Actually, for Japan’s 0.12 billion population, data obtained throughout Japan show 12 and 104 cases of excess mortality in August and October, 2020.

Strict countermeasures taken to forestall the COVID-19 outbreak such as lockdowns and voluntary restrictions against going out induce stress and depress economic activity. They might lower incomes or lead to job loss. These stressors might raise the number of suicides [3]. Actually, excess mortality in suicide was found during the three months of July–September under the first declared state emergency. Those suicide deaths might have occurred even in countries that were more affected than Japan, such as Europe countries or US. However, mortality because of COVID-19 was high in these countries. Therefore the increase in suicide might not be problem that is worth consideration.

Fortunately, morbidity and mortality attributable to COVID-19 were not so high in Japan. Therefore, an increase in suicide might be remarkable only in Japan. A person who dies by suicide is not infected COVID-19, but economic or emotional stress caused by countermeasures against COVID-19 are expected to increase mortality from suicide. Therefore, excess mortality attributable to suicide occurred from COVID-19 outbreak. Countermeasures for COVID-19 should give due consideration to this side effect. In other words, some attempt should be made to minimize the total number of deaths attributable to COVID-19 directly and to limit excess mortality from suicide associated with COVID-19. For the study described herein, we strove to ascertain the optimal level of mobility to minimize overall mortality.

## 2. Method

Direct estimation of the association between mobility and mortality because of COVID-19 requires two steps. First, we identified association among mobility, infectiousness, and the effective reproduction number in an earlier study [4]. Significant association was found between infectiousness and mobility conditional on the temperature. Based on the estimation results, we can predict the infectiousness for a given degree of mobility.

We used mobility data provided by Apple Inc. [5]. The climate data used were the average temperature and relative humidity data for Tokyo during the day [6] from the Japan Meteorological Agency (https://www.data.jma.go.jp/gmd/risk/obsdl/index.php).

If infectiousness can be ascertained, then a simple SIR model [7] can predict the entire outbreak course including the total number of infected patients or duration in outbreak. We assumed the case-fatality rate as 1%.

Excess mortality attributable to suicide was assessed in an earlier study [3]. We regressed the results against average monthly mobility with a lag because it might be presumed that a few months of stress might lead a person to consider suicide. Based on the estimation results, one can estimate the average excess mortality for suicide given a certain condition of mobility. To estimate the total number of suicide deaths for the entire course of the outbreak, we multiplied the average excess mortality from suicide by the outbreak duration.

By following the procedures described above, we obtained the pair of the estimated mortality because of COVID-19 and the expected excess mortality from suicide given a certain value of mobility. We strove to find an optimal degree of mobility for minimization of the overall deaths from these two causes.

## 3. Results

Figure 1 presents mobility in Japan from January 14, 2020 through February 5, 2021. It is apparent that the degree of mobility declined to its lowest of around 60 from late April to mid-May, 2020, when the first emergency state was instituted. The second lowest was in January, 2021, when the second emergency state was instituted.

**Figure 1:**
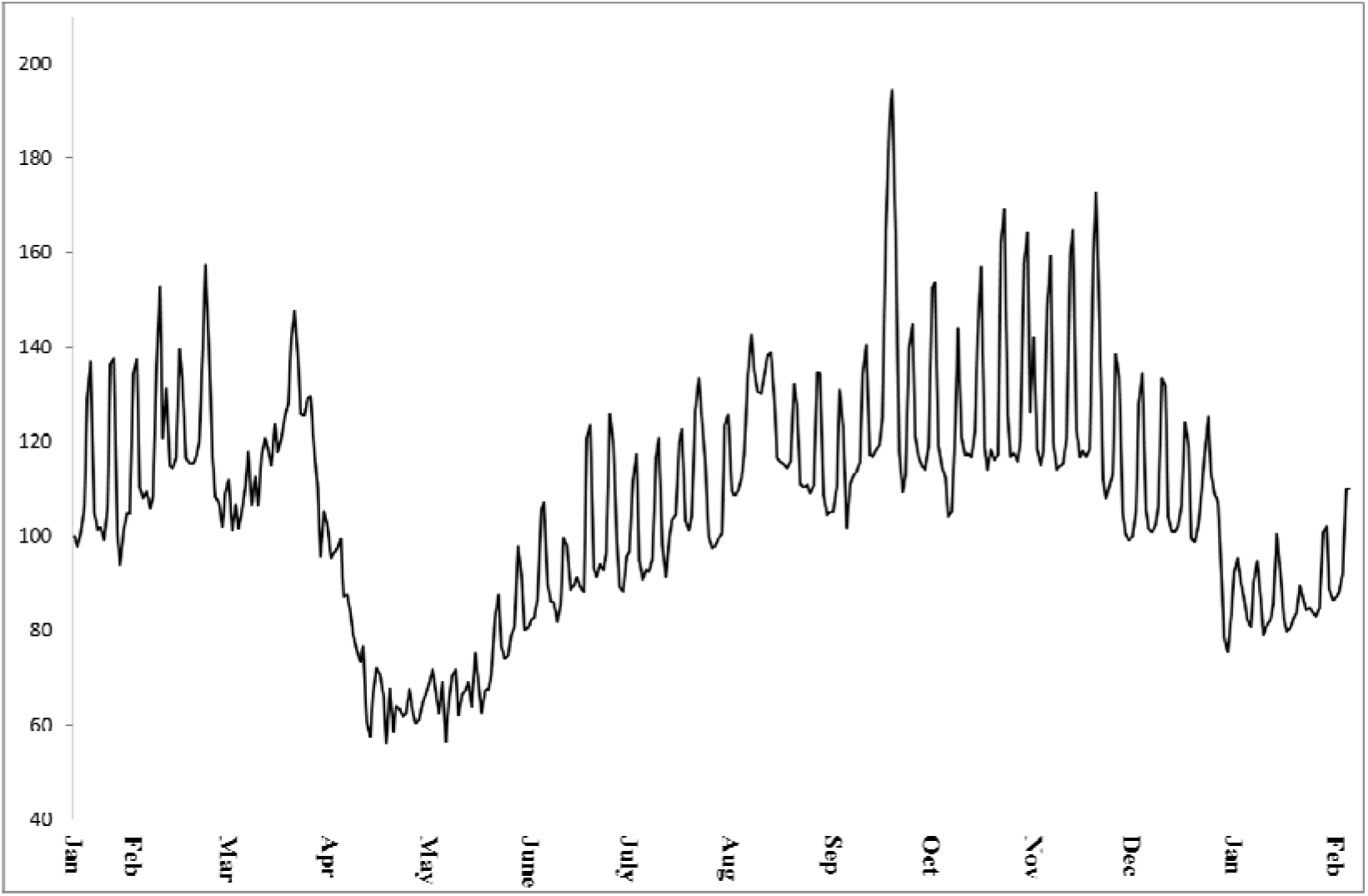
Mobility in Japan from January 14, 2020 to February 5, 2021 Note: Mobilityy data was provided by Apple Inc. The mobility was defined as 100 on January 14, 2020.

Figure 2 depicts the estimated R(*t*) and climate in Japan. It is noteworthy that R(*t*) is shown on the right axis; climate is on the left axis. The latter was normalized to have average of zero and one standard deviation. Results show that R(*t*) was volatile until mid-February because there were very few cases, but it reached the first peak on March 19. Thereafter, it declined to become less than one under the emergency state declaration. After the emergency state declaration, it increased gradually and reached the second peak on June 20. Subsequently, it declined to become smaller than one, but it increased to be higher than one from mid-September. It reached its third peak on October 30. Subsequently it declined, but it was not less than one.

**Figure 2:**
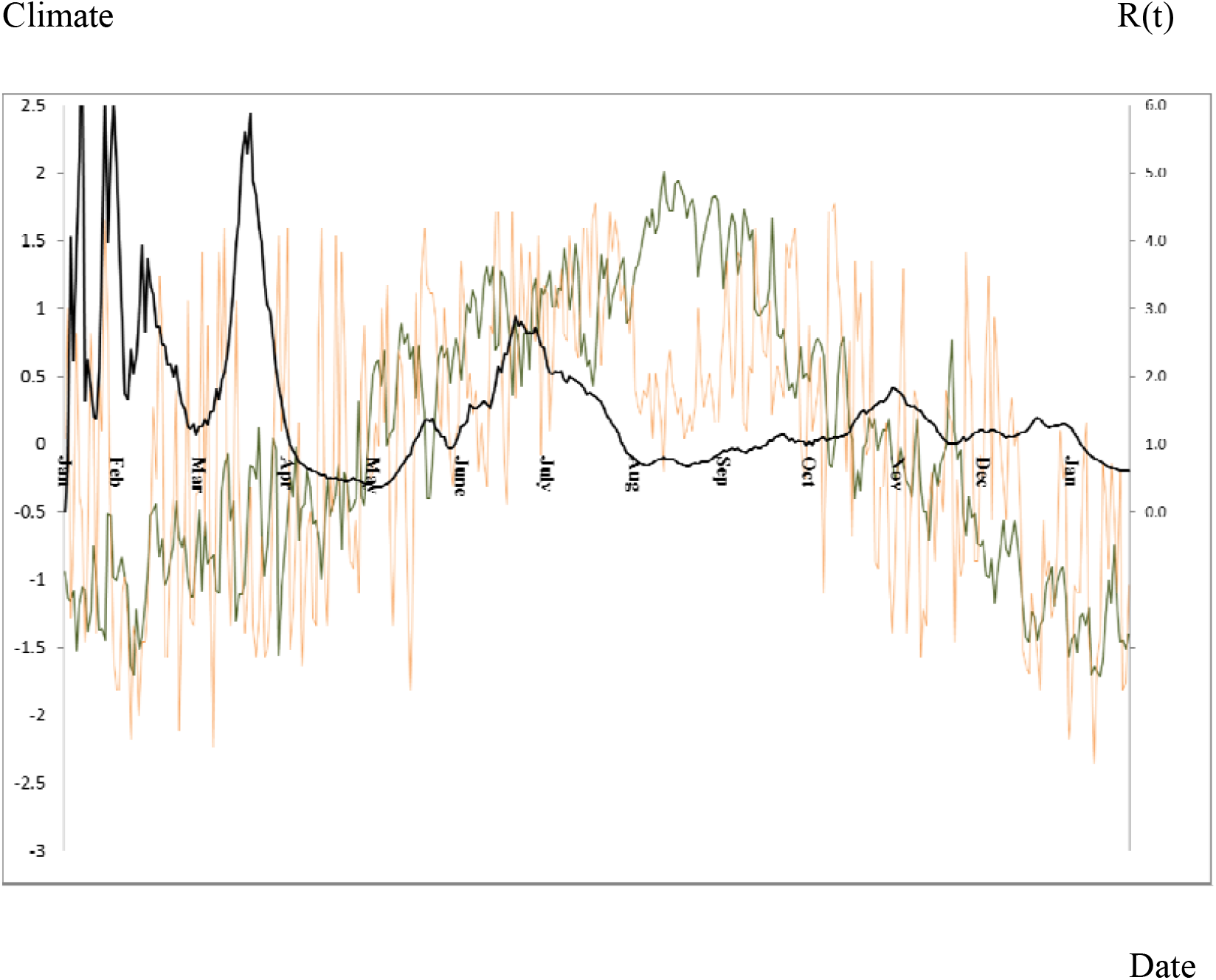
R(t) and climate in Japan. Note: Black line represents R(t). Orange and green lines respectively show humidity and temperature. (t) was measured by the right axis; climate conditions were measured by the right axis. The latter was normalized to be zero average and one standard deviation.

When regressing this information related to climate data and mobility, we obtained

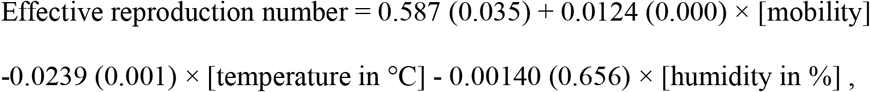

where the number in parenthesis represents a *p*-value. The number of observations was 358. The adjusted coefficient of determination was 0.1381.

Figure 3 depicts excess mortality by suicide found from an earlier study [3]. Among females, large excess mortality was clear in the last three months. Data for males was not greater than among females, but the two highest degrees of excess mortality were shown from 2019. During three months, the total excess mortality attributable to suicide was 810. When regressing this information related to mobility, we obtained the following equation.

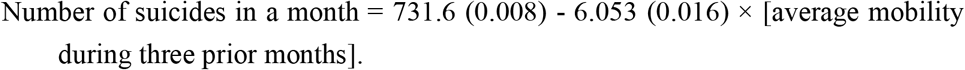

**Figure 3:**
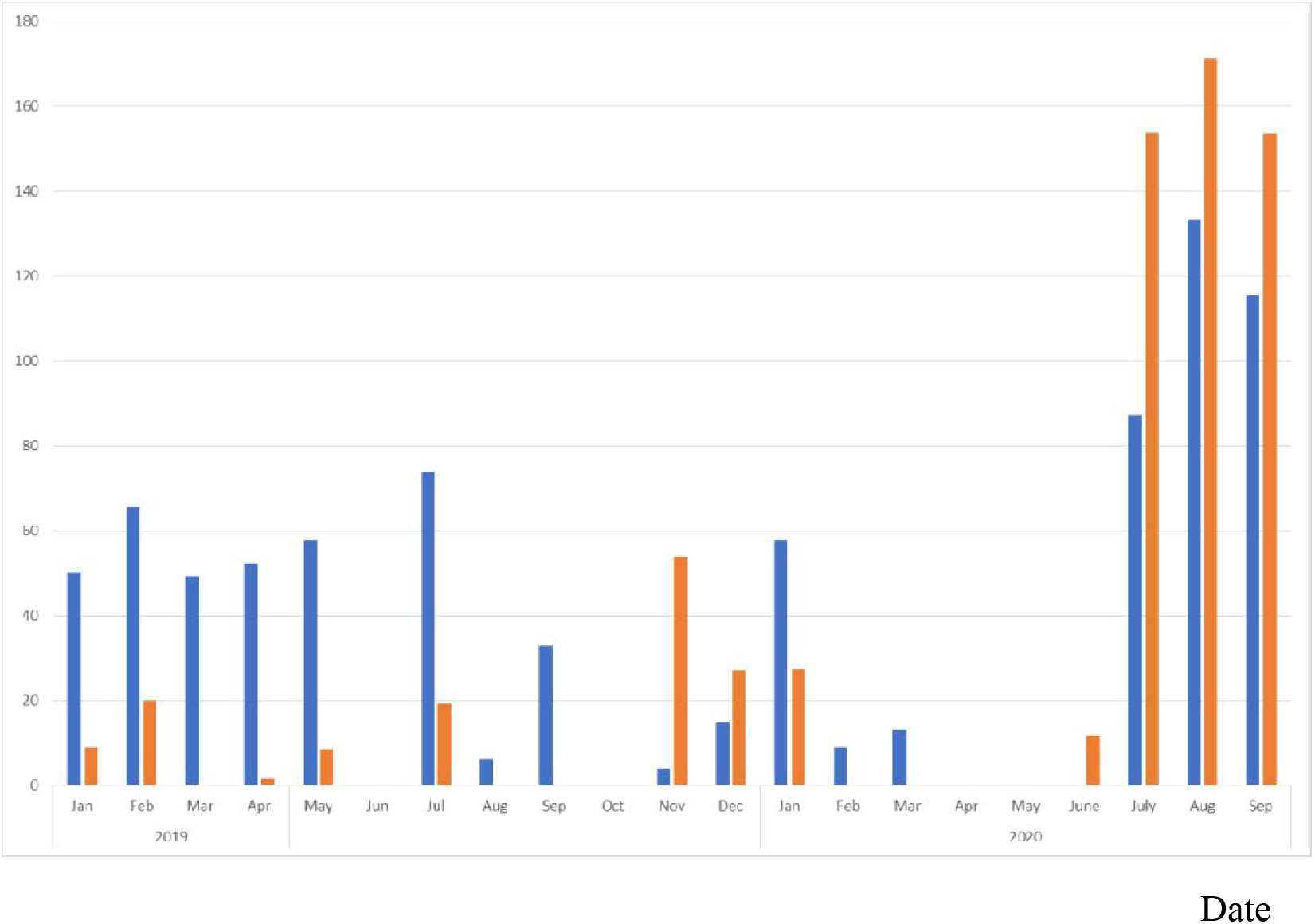
Excess mortality in suicide by gender since 2019 in Japan Date Note: The blue bars represents excess mortaliy in suicide in male, and prange bars indicats the one in fe,ale.

The number of observations was 6. The adjusted coefficient of determination was 0.7514.

Figure 4 shows deaths of these two types by mobility level in the domain of [64, 70] in mobility. The total reached the lowest value when mobility was approximately 65.6, signifying a 33% reduction from the normal level on January 14, 2020.

**Figure 4:**
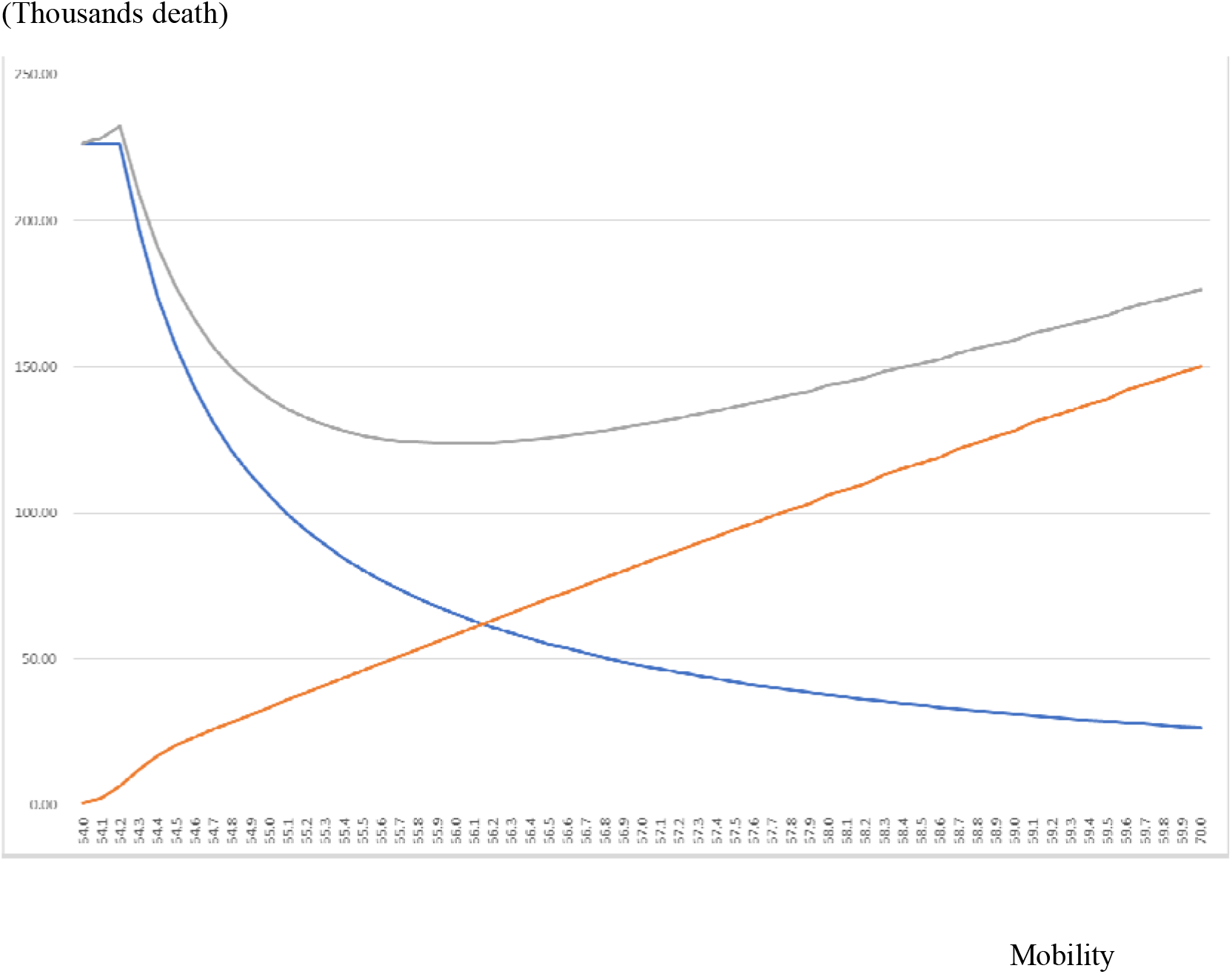
Predicted death due to COVID-19, directly, and excess suicide caused by COVID-19, and sum of them over the entire course of outbreak Note: Green line indicates the predicted death due to COVID-19, directly, blue line indicates excess suicide caused by COVID-19, and sum of them was show as red line. “Mobility” was measured as horizontal axis and it was adjusted as 100 on January 14, 2020, before outbreak.

## 4. Discussion

Results demonstrated that about 34.3% reduction in mobility from the normal level on January 14, 2020 before the outbreak can minimize total deaths caused by COVID-19 infection and suicide caused by the COVID-19 outbreak. This reduction was lower than that achieved when the first emergency state was applied in April and May, 2020. The mobility declined by more than 40% at that time, as shown in Figure 1. However, this target reduction was not achieved thereafter. In other words, reduction was possible, but it was not easy to achieve.

In April, 2020, under the first declared emergency state, 80% reduction in mobility was required [8]. However, it might lead to more than suicide death comparison with COVID-19 death. Therefore, it must be over-restriction in mobility. Actually, it remained as 40% reduction. Some suicide deaths were prevented.

The present study has some limitations. First, our results reflect the situation and experience in Japan in 2020. Consequently, the results cannot be extended to other countries or subsequent years with no modification. International confirmation of our obtained result is anticipated as the next challenge.

Second, although we strove to ascertain a degree of mobility that minimizes the sums of deaths of these two types, if we seek to minimize the total loss of life years, the critical level will rise. In fact, death from COVID-19 occurs directly, and mainly among elderly people. However, suicide attributable to COVID-19 occurs even among younger adults or children as well as among elderly people. Therefore, the loss of life years per mortality case might be greater for suicide from COVID-19 than from COVID-19, directly. Considering differences in life years lost from these two types of death, then the critical level of mobility can be expected to rise.

Third, the higher vaccine coverage hereinafter will raise the critical level of mobility minimizing the total incidence of death. To estimate it, information about vaccine efficacy and its duration in the community are expected to be necessary. Alternatively, because of mutation to higher pathogenicity, the critical level of mobility might be lower. In any case, for future decision making, the critical level of mobility must be re-evaluated according to prevailing circumstances.

## 5. Conclusion

Results demonstrated an optimal level of mobility that minimizes the sum of deaths attributable to COVID-19 infection and suicide caused by the COVID-19 outbreak. Excessive restrictions against mobility raise the number of total deaths.The present study is based on the authors’ opinions: it does not reflect any stance or policy of their professionally affiliated bodies.

## Data Availability

Apple. Mobility trend data.

https://www.apple.com/covid19/mobility

## 6. Acknowledgement

We acknowledge the great efforts of all staff at public health centers, medical institutions, and other facilities who are fighting the spread and destruction associated with COVID-19. Especially, we acknowledge Dr. Nobuhiko Okabe, Kawasaki City Institute for Public Health, Dr.Kiyosu Taniguchi, National Hospital Organization Mie National Hospital, and Dr.Nahoko Shindo, WHO for their helpful support.

## 8. Ethical considerations

All information used for this study was published on the web site of MHLW [1] and Japan Meteorological Agency [7]. Therefore, no ethical issue is presented.

